# Breast cancer epidemiology in women from colombian caribbean region during 2018-2021

**DOI:** 10.1101/2023.08.24.23294568

**Authors:** Valentina Vanessa Pérez Bravo, Valentina Alexandra Vásquez Serge, George Norwood Lobo, Adalgisa Alcocer Olaciregui, Rusvelt Vargas Moranth

**Affiliations:** Universidad Libre, seccional Barranquilla; Universidad del Norte

**Keywords:** Breast cancer, Incidence, Mortality, Prevalence

## Abstract

**Background:** Breast cancer is a global public health problem, representing a significant burden at the personal, collective, economic and quality of life level. It is necessary to consider the incidence, prevalence and mortality of this disease with a regional approach, so the objective of this work is to describe the epidemiological behavior of this tumor in the departments of the Colombian Caribbean Region during the period 2018 to 2021.

**Methodology:** Descriptive, retrospective study. Data from a secondary source (Cuenta de Alto Costo) was taken through the Higia platform. Rates x 100,000 of proportion of new cases reported, prevalence and incidence, by departments are analyzed. Data was imported from a downloadable.csv file into Excel and then into SPSS V22.

**Results:** The indicators analyzed showed a heterogeneous behavior in the 8 departments of the Region. Atlántico and Bolívar were the departments with similar and higher figures in the 4 years observed than those of the country and there were atypical values in San Andrés. La Guajira was the only department with a sustained tendency to increase in incidence, prevalence and mortality.

## INTRODUCTION

The most common neoplasm diagnosed in women is breast cancer, it is a problem in global public health due to its high incidence, prevalence and mortality. According to the Global Cancer Observatory (GLOBOCAN),^1^ in 2020 approximately 2.2 million new cases and 684,996 deaths from this pathology were estimated, making it the second leading cause of death in the world. New cases are predicted to rise over the next few decades to reach nearly 50% higher by 2040.^1^

The region of the Americas accounted for slightly more or less than a quarter of the occurrence of new cases of breast cancer in 2020. In Latin America and the Caribbean there were more than 210,000 new diagnoses of breast cancer, and almost 68,000 deaths.^2^ The proportion of women affected by the disease before the age of 50 (32%) is much higher than in North America (19%).^2^

In Colombia in 2012, 71,442 cases of cancer were diagnosed,^3^ while in 2018 the incidence was 101,893, with an increase of 42.6%.^4^ On the other hand, in 2020 there were a total of cases of cancer of the 113,221 of those 15,509 corresponded to breast cancer, thus representing 13.7% of the Colombian female population. With 4,411 deaths.^1^

According to the World Health Organization (WHO), between 30% and 50% of cancer cases are preventable;^5^ However, cancer continues to be the second cause of death in industrialized countries, after cardiovascular diseases, and was the eighth cause of death in Colombia, in 2019.^6^ This situation is aggravated by the lack of implementation of prevention mechanisms, timely detection and access to treatment.

Breast cancer has shown a downward trend in mortality in developed countries. It is believed that this is related to greater access to health services, which allows early diagnosis and timely treatment of the disease, thus increasing survival.^7^ The condition can occur in women of any social, economic, and ethnic level, although women with greater social disadvantages and fewer resources are the most vulnerable.^8^

The Colombian Caribbean region is made up of 8 departments: Atlántico, Bolívar, Cesar, Córdoba, La Guajira, Magdalena, Sucre and San Andrés, with sociodemographic, cultural and environmental conditions different from the rest of the country, so it is of great importance to analyze the behavior of breast cancer in this area of the nation, in view of the regional approach represented by the new health challenges, for which the present study aims to describe the behavior of three fundamental epidemiological variables to understand population phenomena: incidence, prevalence and mortality from breast cancer in women in the Colombian Caribbean region during the period 2018-2021, the scope of the study is determined by the analysis of the results denoted in the conclusions, in order to guide and project the course of breast cancer. with an action plan by the respective government entities in charge.

## MATERIALS AND METHODS

A descriptive and retrospective study was carried out, in which data from a secondary source was taken: Cuenta de Alto Costo (CAC).^9^ Through the “Higia” Portal, in which the relevant training was previously accessed to obtain user and password for it. The demographic and morbidity and mortality dashboard corresponding to cancer was entered and once there, year by year (2018 to 2021) and by department (Atlántico, Bolívar, Cesar, Córdoba, La Guajira, Magdalena, San Andrés and Sucre), the data were downloaded. Data of:

- PCNR (Proportion of new cases reported): number of cases of invasive breast cancer with a date of oncological diagnosis.
- Prevalence: total number of cases diagnosed with invasive breast cancer.
- Mortality: number of deaths from invasive breast cancer.

Each of the three indicators are expressed as: number of cases per 100,000 inhabitants/members, within each reported year.

The data were downloaded as a flat.csv file, which was imported from Microsoft Excel and subsequently analyzed using SPSS Version 22. Both the PCNR, as well as the prevalence and mortality were conceived as rates standardized by the direct method, globally and as crude rates when discriminating the data by age (from 30 to 79 years, by five-year groups).

## RESULTS

The lowest PCNR x 100,000 occurred in the department of La Guajira in 2018, with 5.25, followed by San Andrés in 2019 with 6.46, while the highest values were in the latter department, but in 2018, with 42.59 and in Bolívar in 2020 with 28.76. In most departments these rates presented a fluctuating value, but in La Guajira a clear progressive trend towards increase was observed, going from 5.25 in 2018 to 20.10 in 2021; This growing pattern is observed, but only from 2019 to 2021 in Atlántico and Cesar and from 2018 to 2010 in Sucre (Graph 1).

**Graph 1.**
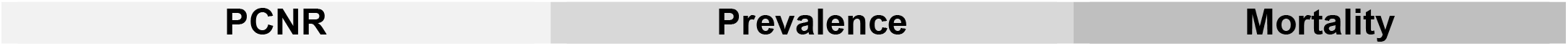
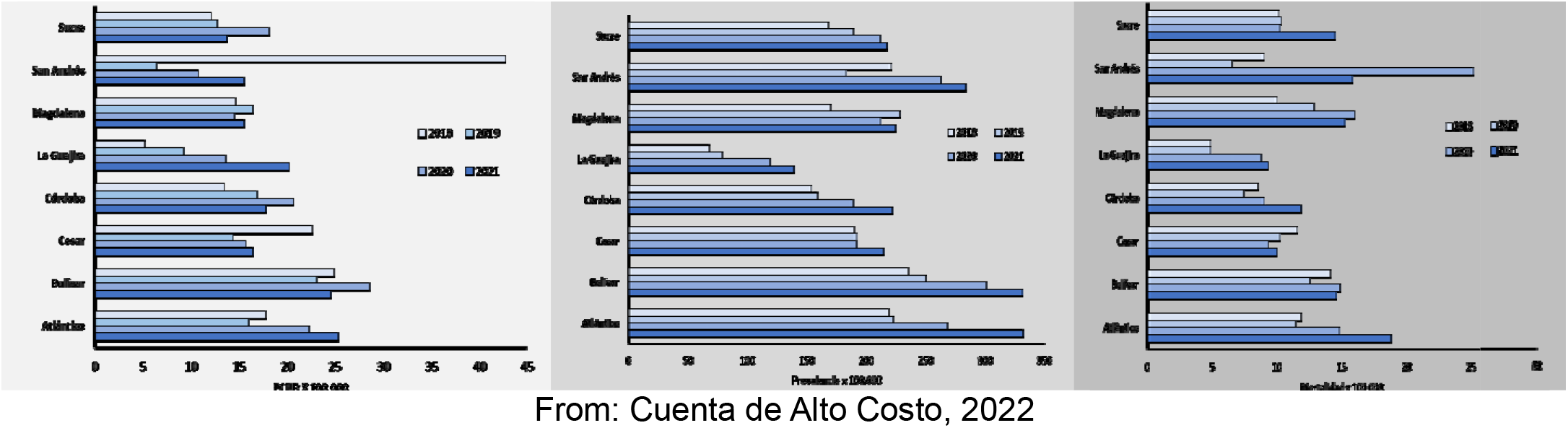
PCNR, Prevalence and Mortality x 100,000, for invasive breast cancer in the departments of the Colombian Caribbean Region, during 2018 to 2021.

By age, the crude PCNR rates showed a trend as age increased, from 30 to 79 years, in almost all departments, with the exception of La Guajira, where the maximum peaks were in the 65 to 69 age group and in terms of In general, in Bolívar in the year 2020 the highest rate was observed for the group of 70 to 74 years with 111.9 × 100,000 (Graph 2) For the department of San Andrés, the values by age are not shown, being quite scattered and with many values equal to zero (Annex 1).

**Graph 2.**
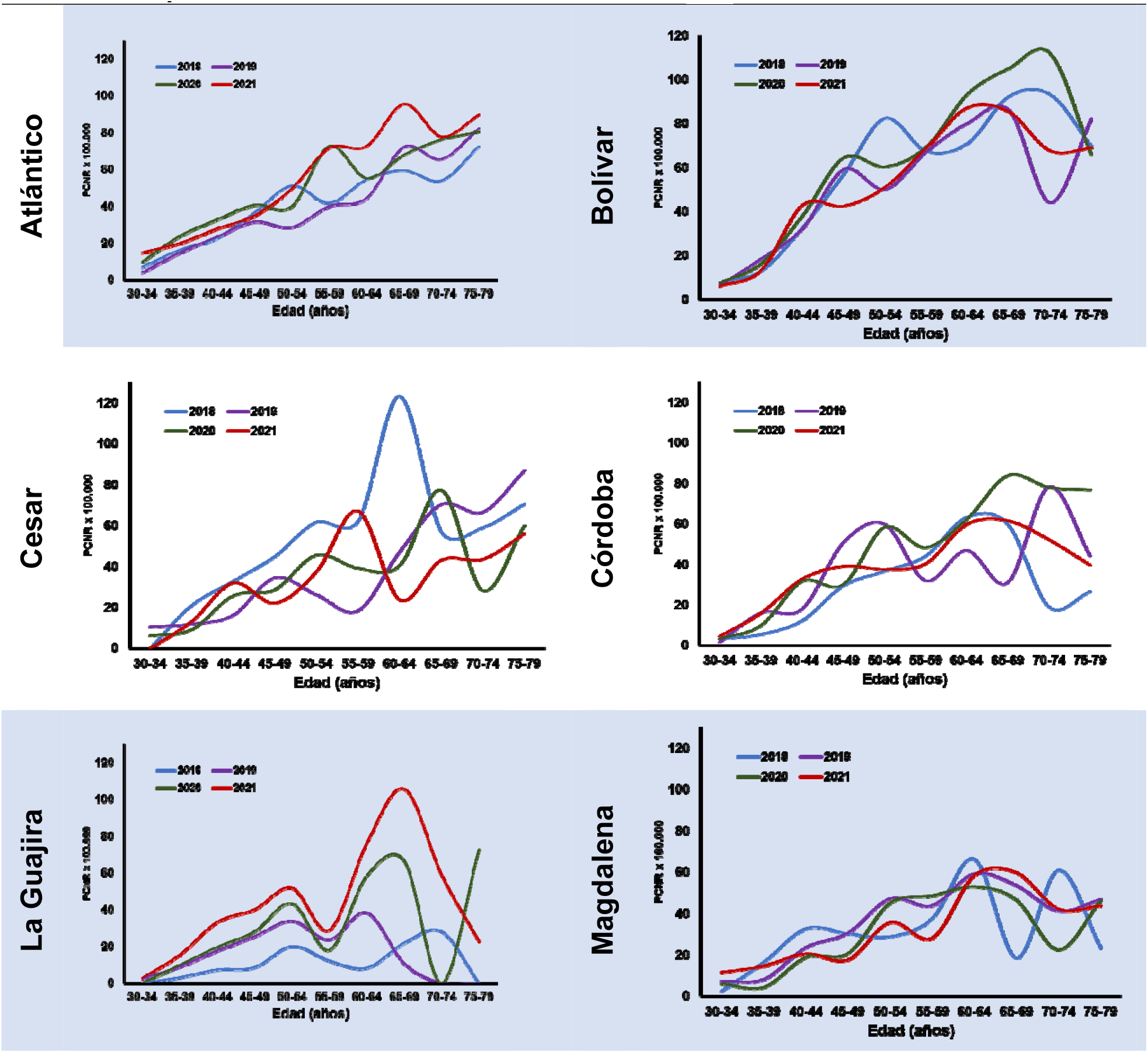
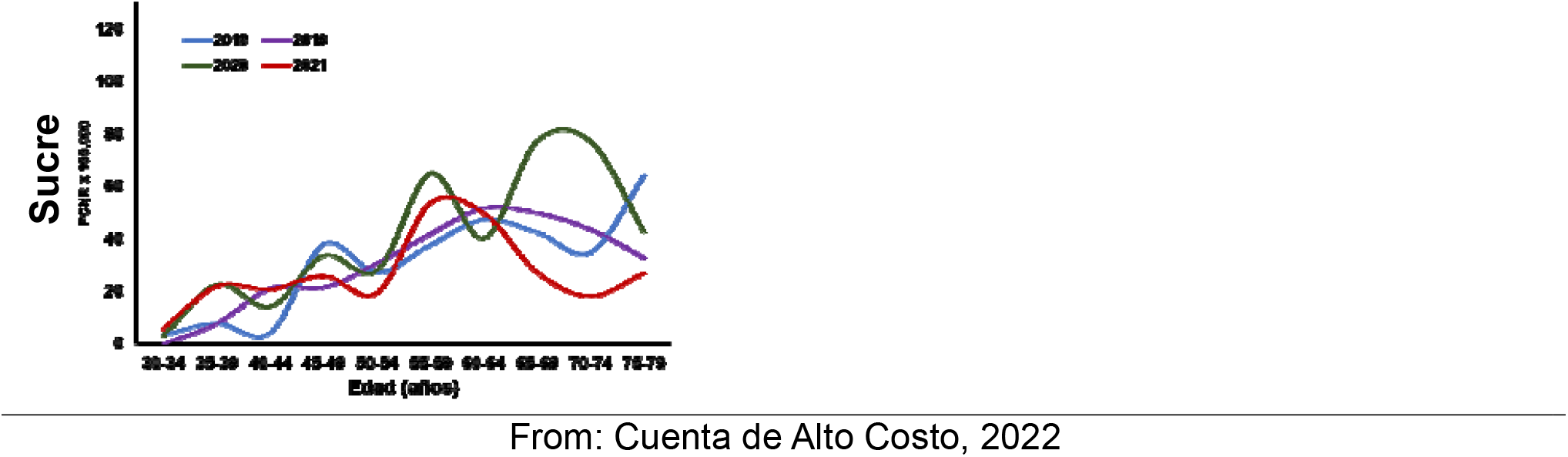
Crude rates of PCNR x 100,000 by age, of invasive breast cancer, in the departments of the Colombian Caribbean Region during 2018 to 2021.

With the exception of San Andrés and Magdalena, in all the departments studied there was an upward trend in the value of the prevalence of invasive breast cancer, and the highest rates were in Atlántico and Bolívar: close to 330 × 100,000 in 2021 (Graph 1). Regarding mortality, there was a slight downward trend in Cesar, going from 11.55 × 100,000 to 9.93, from 2018 to 2021. Atlántico and Magdalena had the highest rates in almost every year, but in 2020 an apparently atypical figure was presented in San Andrés: 25.16 (Graph 1).

## DISCUSSION

Values higher than the mortality reported by the IARC for Colombia1 were found in the year 2020: 13.1 × 100,000, in each of the years selected for the departments of Atlántico and Bolívar, reaching a value of 18.76 in the first in 2021, while, for San Andrés, the mortality rate was almost double the national one. The incidence and prevalence are not comparable to those of the IARC, given the different methodologies used for their calculation, but when compared with the CAC data,9 which range from 18.7 to 25.8 during the 4 years of observation, the rates were higher in the department of Bolívar and very similar for Atlántico, while the national prevalences (from 230.5 to 306.7) were also lower than those of Bolívar and similar to those of Atlántico; for the rest of the departments, the PCNR and prevalences were clearly lower than those of the country.

As a limitation, there is the retrospective nature and secondary source of this study, but the methodology consolidated by CAC for the calculations is accepted as official by the Ministry of Health and Social Protection, and although there seem to be underreporting in some years for certain departments with San Andrés, it is expected that subsequent adjustments will be carried out by CAC.

## CONCLUSIONS

Breast cancer in the departments of the Colombian Caribbean Region presents comparable epidemiological indicators with national data. There was no homogeneous pattern in the 8 departments, but it was found that Atlántico and Bolívar have the highest incidences, prevalences, and mortality globally in the 4 years of observation and that for Bolívar the values are higher than those reported for the country.

## Data Availability

All data produced in the present study are available upon reasonable request to the authors.

