## Supplemental Tables for "Breast cancer epidemiology in women from colombian caribbean region during 2018-2021"

### ANNEX 1. RESULT CHARTS

**Chart 1. Rates x 100,000 of Incidence (PCNR), Prevalence and Mortality due to invasive breast cancer in departments from the Colombian Caribbean Region during 2018 to 2021**

| | Años | Atlántico | Bolívar | Cesar | Córdoba | La Guajira | Magdalena | San Andrés | Sucre |
|---|---|---|---|---|---|---|---|---|---|
| Incidence | 2021 | 25,21 | 24,56 | 16,37 | 17,81 | 20,19 | 15,49 | 15,57 | 13,80 |
| | 2020 | 22,20 | 28,67 | 15,69 | 20,55 | 13,62 | 14,48 | 10,75 | 18,11 |
| | 2019 | 15,95 | 23,10 | 14,29 | 16,82 | 9,26 | 16,36 | 6,46 | 12,72 |
| | 2018 | 17,75 | 24,76 | 22,50 | 13,42 | 5,25 | 14,62 | 42,59 | 12,05 |
| Prevalence | | Atlántico | Bolívar | Cesar | Córdoba | La Guajira | Magdalena | San Andrés | Sucre |
| | 2021 | 332,48 | 331,36 | 214,55 | 221,43 | 138,79 | 223,61 | 283,43 | 216,77 |
| | 2020 | 267,89 | 301,09 | 191,33 | 188,71 | 118,58 | 212,49 | 262,21 | 212,24 |
| | 2019 | 223,44 | 250,16 | 191,12 | 158,22 | 78,35 | 227,95 | 183,28 | 189,30 |
| | 2018 | 219,38 | 235,31 | 190,17 | 154,34 | 68,00 | 170,16 | 220,01 | 167,61 |
| Mortality | | Atlántico | Bolívar | Cesar | Córdoba | La Guajira | Magdalena | San Andrés | Sucre |
| | 2021 | 18,76 | 14,62 | 9,93 | 11,89 | 9,32 | 15,26 | 15,81 | 14,43 |
| | 2020 | 14,75 | 14,86 | 9,30 | 9,01 | 8,80 | 15,96 | 25,16 | 10,27 |
| | 2019 | 11,42 | 12,54 | 10,26 | 7,42 | 4,88 | 12,82 | 6,51 | 10,34 |
| | 2018 | 11,92 | 14,06 | 11,55 | 8,49 | 4,89 | 9,97 | 9,02 | 10,09 |

From: Cuenta de Alto Costo, 2022

**Tabla 2. Crude incidence rates (PCNR) by age of invasive breast cancer in departments from the Colombian Caribbean Region during 2018 to 2021.**

| Age | Atlántico | | | | Bolívar | | | | Cesar | | | | Córdoba | | | |
|---|---|---|---|---|---|---|---|---|---|---|---|---|---|---|---|---|
| | 2018 | 2019 | 2020 | 2021 | 2018 | 2019 | 2020 | 2021 | 2018 | 2019 | 2020 | 2021 | 2018 | 2019 | 2020 | 2021 |
| 30-34 | 6,92 | 3,96 | 9,71 | 14,87 | 7,83 | 6,44 | 7,57 | 6,11 | 0,00 | 10,46 | 6,21 | 0,00 | 3,19 | 1,55 | 3,08 | 4,56 |
| 35-39 | 16,09 | 14,63 | 23,63 | 19,87 | 13,13 | 18,59 | 16,42 | 13,30 | 20,99 | 11,86 | 9,15 | 13,18 | 5,42 | 15,94 | 9,70 | 16,02 |
| 40-44 | 22,64 | 23,24 | 32,70 | 27,87 | 32,76 | 32,27 | 38,27 | 43,01 | 32,89 | 16,20 | 25,69 | 32,09 | 12,20 | 17,92 | 31,56 | 32,81 |
| 45-49 | 36,44 | 31,54 | 40,60 | 34,87 | 57,20 | 59,27 | 64,58 | 42,48 | 44,93 | 34,53 | 28,71 | 22,29 | 29,41 | 50,68 | 29,91 | 39,08 |
| 50-54 | 51,08 | 28,56 | 39,93 | 49,58 | 82,49 | 50,14 | 60,41 | 51,03 | 61,80 | 26,21 | 45,56 | 37,56 | 36,61 | 59,97 | 58,30 | 37,64 |
| 55-59 | 41,69 | 39,99 | 72,56 | 71,61 | 67,44 | 67,22 | 69,44 | 68,89 | 62,15 | 18,37 | 39,20 | 67,24 | 44,11 | 31,98 | 48,31 | 40,36 |
| 60-64 | 54,50 | 44,32 | 55,05 | 72,64 | 71,10 | 80,18 | 93,54 | 87,30 | 123,16 | 47,13 | 40,45 | 23,95 | 63,24 | 46,95 | 62,20 | 59,89 |
| 65-69 | 59,46 | 72,18 | 67,83 | 95,56 | 92,42 | 86,10 | 105,14 | 85,27 | 57,15 | 70,35 | 77,44 | 43,22 | 59,78 | 31,03 | 83,95 | 61,69 |
| 70-74 | 53,89 | 65,46 | 76,05 | 77,82 | 93,31 | 44,11 | 111,91 | 67,64 | 59,00 | 66,56 | 28,28 | 43,67 | 19,04 | 78,39 | 78,03 | 52,34 |
| 75-79 | 72,49 | 82,25 | 80,24 | 89,62 | 69,81 | 82,29 | 65,84 | 68,91 | 70,50 | 86,79 | 59,86 | 56,02 | 26,53 | 44,13 | 76,82 | 39,75 |

| Age | La Guajira | | | | Magdalena | | | | San Andrés | | | | Sucre | | | |
|---|---|---|---|---|---|---|---|---|---|---|---|---|---|---|---|---|
| | 2018 | 2019 | 2020 | 2021 | 2018 | 2019 | 2020 | 2021 | 2018 | 2019 | 2020 | 2021 | 2018 | 2019 | 2020 | 2021 |
| 30-34 | 0,00 | 2,46 | 0,00 | 2,71 | 2,48 | 7,32 | 6,01 | 11,59 | 0,00 | 0,00 | 39,81 | 0,00 | 3,43 | 0,00 | 3,01 | 5,82 |
| 35-39 | 3,06 | 8,92 | 9,76 | 15,40 | 16,48 | 8,21 | 4,31 | 14,64 | 40,68 | 0,00 | 0,00 | 0,00 | 7,85 | 7,77 | 22,67 | 22,10 |
| 40-44 | 7,37 | 17,53 | 19,53 | 33,11 | 32,69 | 23,56 | 18,86 | 20,59 | 0,00 | 0,00 | 47,19 | 46,13 | 4,31 | 21,19 | 14,24 | 20,77 |
| 45-49 | 8,60 | 25,35 | 28,04 | 40,02 | 30,25 | 30,56 | 20,70 | 17,70 | 80,68 | 0,00 | 0,00 | 93,33 | 38,06 | 21,59 | 33,70 | 25,98 |
| 50-54 | 20,12 | 33,77 | 43,24 | 51,62 | 28,77 | 47,16 | 45,14 | 35,76 | 0,00 | 69,76 | 41,25 | 0,00 | 27,15 | 30,68 | 28,04 | 19,58 |
| 55-59 | 12,19 | 23,64 | 18,04 | 28,91 | 37,70 | 43,83 | 48,66 | 28,16 | 153,61 | 36,83 | 41,93 | 40,68 | 37,75 | 42,09 | 65,06 | 53,86 |
| 60-64 | 8,26 | 38,28 | 58,29 | 76,01 | 66,28 | 59,20 | 53,05 | 58,27 | 207,47 | 0,00 | 0,00 | 106,50 | 47,31 | 51,83 | 40,32 | 49,79 |
| 65-69 | 21,83 | 10,68 | 66,42 | 105,46 | 18,54 | 53,41 | 46,60 | 59,67 | 80,06 | 0,00 | 0,00 | 0,00 | 42,51 | 49,97 | 77,46 | 27,31 |
| 70-74 | 28,08 | 0,00 | 0,00 | 58,77 | 61,01 | 41,17 | 22,55 | 42,03 | 268,10 | 0,00 | 0,00 | 0,00 | 34,87 | 43,71 | 77,19 | 18,20 |
| 75-79 | 0,00 | 0,00 | 72,45 | 22,72 | 23,33 | 46,82 | 46,27 | 43,72 | 0,00 | 0,00 | 0,00 | 0,00 | 64,07 | 32,70 | 42,69 | 27,11 |

From: Cuenta de Alto Costo, 2022
